# Whole-genome sequencing in mitral valve prolapse – an exploratory study on genes associated with mitral valve prolapse and cardiac arrhythmias

**DOI:** 10.64898/2026.09.03.26362147

**Authors:** Nikhil Arora, Julie Bergh, Christine Rootwelt-Norberg, Christian Five, Bjørn Olav Åsvold, Brooke N Wolford, Cecilie Bugge, Anna Isotta Castrini, Nina Eide Hasselberg, John-Peder Escobar Kvitting, Eivind Coward, Kristian Hveem, Kristina Hermann Haugaa

## Abstract

**Background:** Mitral valve prolapse (MVP) is a leading cause of mitral regurgitation in high-income countries. Although the course of MVP is often benign, a subset of patients progresses to significant valvular insufficiency, and a smaller subset develops ventricular arrhythmias or sudden cardiac death. This study aimed to identify genetic variants associated with MVP and/or mitral annulus disjunction (MAD) using whole-genome sequencing (WGS) in a Norwegian cohort.

**Methods:** WGS was performed in 93 individuals diagnosed with MVP and/or MAD, of whom 92 passed genomic quality control criteria. Variants were called using Genome Analysis ToolKit (GATK) Best Practices and compared with non-Finnish European reference data from gnomAD (n = 34 029). Analyses were restricted to curated gene panels comprising: (i) non-syndromic MVP and cardiomyopathy genes, (ii) syndromic/connective tissue disorder genes, and (iii) arrhythmia-related genes. Enriched or underrepresented variants were identified using Fisher’s exact test with Bonferroni correction for multiple testing. Independent variants were derived through linkage disequilibrium pruning. A sensitivity analysis was conducted using low-coverage WGS data from an independent large Norwegian cohort – the Trøndelag Health Study (HUNT).

**Results:** Across predefined gene panels, several coding and regulatory variants were significantly associated with MVP and/or MAD. A missense variant in *FLNA* and regulatory variants in *TBX5* and *SMAD4* were enriched in these patients. Additional significant associations involved regulatory variants in *XYLT1, HS3ST4, SH3PXD2B*, and a synonymous variant in *COL1A2*. Furthermore, several regulatory variants in arrhythmia-related genes (*KCNK3, CACNA1D, NR2F1, KCNJ4* and others) were identified. Sensitivity analyses revealed population-level differences, underscoring the need of cautious interpretation of these findings.

**Conclusion:** This targeted WGS study expands the genetic landscape of MVP and/or MAD, identifying several coding and regulatory variants across extracellular matrix, developmental, and electrophysiological pathways. These results support a multifactorial genetic architecture of MVP.

**Graphical Abstract:** Whole-genome sequencing of 92 Norwegian patients with mitral valve prolapse (MVP) and/or mitral annulus disjunction (MAD) identified coding and regulatory genetic variants across three curated gene categories: (i) non-syndromic MVP/cardiomyopathy genes, (ii) syndromic MVP/connective tissue disorder genes, and (iii) arrhythmia-related genes. Key associations included variants in *FLNA, TBX5, COL1A2, SMAD4*, and several electrophysiological genes including *KCNK3, CACNA1D, NR2F1*, and *KCNJ4*. The findings implicate extracellular matrix (ECM) remodeling, transforming growth factor-β (TGF-β) signaling, cardiac developmental pathways, cytoskeletal integrity, and electrophysiological mechanisms, supporting a multifactorial genetic architecture of MVP/MAD.

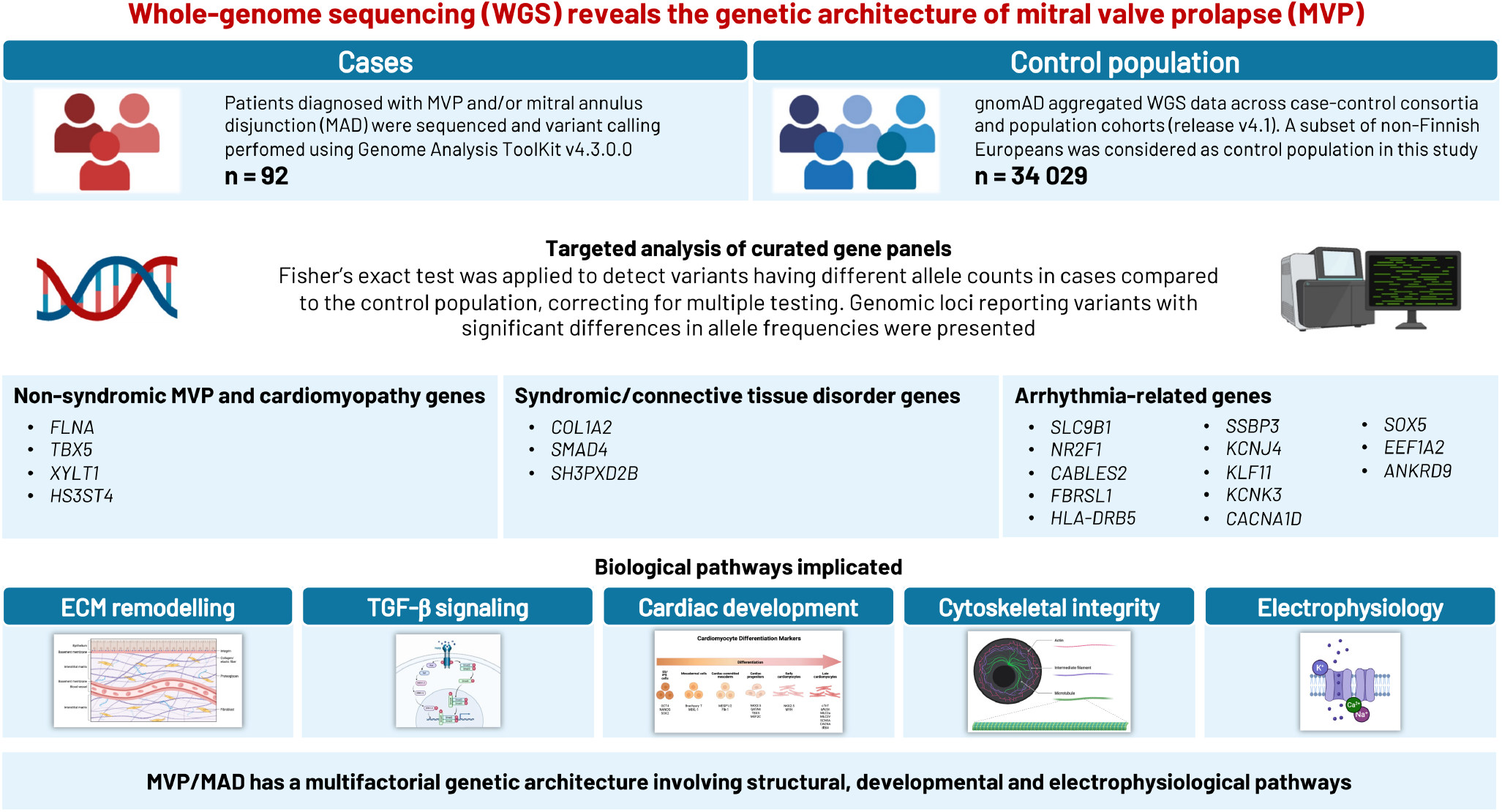

## Introduction

Mitral valve prolapse (MVP) is a valvular heart disease affecting 2-3% of the population^1^, and constitutes a leading cause of mitral regurgitation (MR) in high-income countries^2^. While many individuals remain asymptomatic, a subset develops progressive valvular insufficiency that may lead to adverse outcomes, including heart failure, atrial fibrillation, and importantly an increased risk of life-threatening ventricular arrythmias (VAs) and sudden cardiac death (SCD)^3–6^.

The genetic basis of MVP is supported by familial clustering^7,8^, with 35-50% of cases exhibiting a familial pattern^9^, consistent with a substantial heritable component. While MVP predominantly manifests sporadically, it is also frequently present in a number of syndromes and systemic collagen disorders, such as pseudoxanthoma elasticum, Marfan syndrome, Loeys-Dietz syndrome, Ehlers-Danlos syndrome and osteogenesis imperfecta^10–15^. This suggests a central role of connective tissue abnormalities in the pathophysiology of MVP. Numerous studies have shown associations between MVP and various genetic loci^16–21^. These genes are delineated into those causing syndromic forms of MVP and those involved in non-syndromic presentations, encompassing sporadic and familial cases^22^. The roles of several genes have been thoroughly identified in the syndromic forms of MVP, while in the latter a progressively increasing number of candidate genetic loci have been thoroughly investigated.

Non-syndromic MVP is regarded as a polygenic disorder arising from alterations in multiple genetic pathways, with several genetic variants contributing to disease susceptibility^21^. Certain disease-causing mutations leading to non-syndromic MVP have been identified in genes such as *PLD1*^23^, *DCHS1*^24^, *DZIP1*^25^, and *FLNA*^26,27^. Previously, familial studies and linkage analysis in large MVP families identified loci associated with MVP on chromosomes 16p11-p12 (*MMVP1*), 11p15.4 (*MMVP2*), 13q31-32 (*MMVP3*), and Xq28 (X-linked MVP), with *MMVP2* now identified as *DCHS1* gene, *MMVP3* as *DZIP1* gene, and X-linked MVP as *FLNA* gene^19,25^. These genes are involved in pathways governing membrane integrity, cell signaling, cell adhesion, cytoskeletal integrity, and ciliary function^23–27^. Mutations in *FLNC*, encoding filamin C – an actin-binding protein vital for cytoskeleton, have also been reported as a monogenic cause of MVP^28^. More recently, genes commonly associated with cardiomyopathies (e.g., *DSP, TTN, MYH6, HCN4, LMNA*)^18,29^, as well as multiple genetic loci identified by genome-wide association studies (GWAS) like *LMCD1, TNS1*, and *MSRA*, implicate a polygenic architecture of MVP^16,30^. Notably, a recent GWAS meta-analysis including 4884 cases of MVP and 434 649 controls observed an overall SNP-based heritability of 0.22 and highlighted novel loci in genes such as *LTBP2, TGFB2, ALPK3, BAG3, RBM20*, and *SPTBN1* – emphasizing the roles of TGFβ signaling, cytoskeleton integrity, and cardiomyopathy pathways in MVP pathogenesis^21^.

Cardiac arrhythmias, both ventricular and supraventricular forms, may present as complications of MVP^2,31^. A distinct subtype – termed arrhythmogenic MVP (AMVP) – has recently been defined^32^. However, the coexistence of MVP and SCD remains to be firmly established^33^. A recent study found an association between MVP with VAs and SCD independent of valve incompetence and heart failure^34^, suggesting these patients have an increased arrhythmic risk regardless of the degree of hemodynamic and myocardial dysfunction. Furthermore, the mechanisms underlying arrhythmogenesis in MVP remain poorly understood, with competing hypotheses implicating either – a heritable proarrhythmic genetic substrate may underlie MVP, fibrotic left ventricular remodeling in MVP as the primary driver of arrhythmogenesis, or an interaction between these factors.

Despite the studies contributing to extend knowledge on pathogenesis of MVP, the genetic etiology of MVP and their relationship to arrhythmia risk remain elusive. In this study, we sought to identify novel genetic variants associated with MVP in a previously unstudied population. We hypothesized that several genetic variants with plausible causal links to MVP could be detected, thereby delineating its genetic basis and addressing to bridge knowledge gaps in understanding its underlying pathophysiology.

## Methods

### Study population

Patients with MVP and/or mitral annulus disjunction (MAD) were recruited from two centers – Oslo University Hospital and Drammen Hospital, as previously reported^35^. All patients underwent an evaluation at the Department of Cardiology, Oslo University Hospital, Rikshospitalet during the period 2014 to 2017. Blood samples for genetic analyses were collected from 96 of the patients, and 93 of these patient’s samples (100% European ancestry) were available for the final analysis. This study was conducted in accordance with the Declaration of Helsinki and was approved by the Regional Committee for Medical Research Ethics (Project number: 2015/596/REK North). All the study patients provided written informed consent.

#### Clinical characteristics

MVP was defined as systolic superior displacement ≥2 mm of any part of the mitral leaflet into the left atrium detected on echocardiography in the parasternal long-axis view^32^. MAD was defined as a ≥1 mm separation of the left atrial wall junction of the posterior mitral leaflet and the ventricular myocardium during end systole detected on echocardiography in the parasternal long-axis view or apical four-chamber view^35^. Cardiac volumes and functions were assessed and mitral regurgitation was graded according to the guidelines using echocardiography^36–38^. Data from all patients have been reported in previous studies^35,39^.

#### Whole-genome sequencing (WGS)

All genomic DNA samples were extracted from peripheral blood using standard procedures at the Trøndelag Health Study (HUNT) Biobank^40^. WGS (30x) was carried out at the Norwegian Sequencing Centre – Oslo University Hospital. Data preprocessing involved genomic quality control checks using fastQC on the sampled reads. Reads were mapped against the human reference genome (GRCh38/hg38) using Burrows-Wheeler Aligner^41^. One sample did not pass the quality control checks and was excluded. To this end, analysis-ready reads files in BAM format were generated, which were consequently used as an input for the next step, known as variant calling (or variant discovery).

#### Data analysis and filtering

Genome Analysis ToolKit (GATK) release v4.3.0.0 was used for calling variants, filtering, and evaluation^42^. We used the GATK Best Practices, which provide a comprehensive guide for performing variant discovery analysis on high-throughput sequencing data^43^. We began by calling variants for each sample using the GATK HaplotypeCaller tool on the per-sample BAM files, generating per-sample GVCF files. These GVCF files from multiple samples were then combined and subjected to joint variant calling using the GATK GenomicsDBImport and GATK GenotypeGVCFs tools, respectively. This process generated raw single nucleotide polymorphism (SNPs) and short insertions and deletions (indels) files in VCF format, which were subsequently used for filtering and annotation.

Raw SNPs and indels variants were filtered to ensure high confidence variant calls. This study employed hard filtering method using the GATK VariantFilteration, GATK VariantRecalibration, and GATK ApplyVQSR tools. The filtration criteria as specified elsewhere^42^, considered a QualByDepth (QD) filter threshold of less than 2.0, a FisherStrand (FS) filter threshold greater than 60, and a StrandOddRatio (SOR) filter threshold greater than 4.0. In addition, genotype filtration (sample-level) was performed with a depth (DP) filter threshold of less than 10 and a genotype quality (GQ) filter threshold of less than 20. After completing the filtration process, variants passed were extracted, resulting in analysis-ready variant files. These files were annotated using ANNOVAR v2024Feb19^44^.

### Controls selection

The Genome Aggregation Database (gnomAD) is the largest publicly available human genome allele frequency reference dataset^45^. gnomAD release v4.1 aggregated whole-genome sequence data from 153 030 individuals across various case-control consortia and population cohorts, releasing a highquality call set of 76 215 individuals^45,46^. Frequency information was available for several strata of this dataset based on attributes such as ancestry and sex for each of 644 267 978 short nuclear variants. We used data from the non-Finnish European genetic ancestry group (n = 34 029) as control population for this study.

### Candidate gene selection

To inform genetic variant selection, candidate gene identification was guided by recent reviews on MVP, latest GWAS studies, key primary studies and by screening reference lists of relevant articles to collate a list of genes implicated in MVP. This entailed compiling genes associated with non-syndromic MVP, genes jointly linked to MVP and cardiomyopathy phenotypes, and genes with more limited or emerging evidence of association with MVP (more information in Supplementary file, Table S1). To investigate potential connective tissue pathology underlying MVP, genes known to be involved in syndromic forms of MVP and associated systemic collagenopathies were guided by recent reviews on MVP (more information in Supplementary file, Table S2). Furthermore, to explore the hypothesis of a heritable proarrhythmic genetic substrate in MVP, genes previously established as being associated with cardiac arrhythmias irrespective of valve pathology were guided by lastest GWAS studies on phenotypes linked with conduction anomalies (more information in Supplementary file, Table S3). The resulting gene lists served as the foundation for subsequent genetic analyses in the present study.

### Statistical analysis

All genetic variants in genetic regions previously known to be associated with non-syndromic MVP, MVP and cardiomyopathies along with genes having lesser-known associations with MVP (Supplementary file 1, Table S1) were identified from our sample. Variants were extracted from annotated VCF file using the genes of interest as query terms. Therefore, all variants assigned to the corresponding genes by the annotation pipeline were evaluated, including exonic, intronic, untranslated region (UTR), upstream, downstream, and intergenic variants. Allele counts were calculated for these genetic variants using their sequenced information from each sample. Fisher’s exact test was applied to detect enriched and underrepresented genetic variants, with Bonferroni correction for the number of genetic variants used to account for multiple testing. In this analysis, the allele counts for genetic variants in our sample (n = 92) were compared with allele counts for non-Finnish European genetic ancestry group (n = 34 029) as the controls. Furthermore, pruning was performed using *SNPclip* function of *LDlinkR* package in R, to remove genetic variants in high linkage disequilibrium (LD) and identify a set of independent genetic variants^47^. The 1000 Genome Project European sub-population was used as a reference, with R^2^ threshold of 0.2 and minor allele frequency (MAF) threshold of 0.01 for LD pruning.

In addition, genetic variants in genes involved in syndromic forms of MVP and associated systemic collagenopathies (Supplementary file, Table S2), and genetic variants in genes previously established as contributing to cardiac arrythmias (Supplementary file, Table S3) were identified from our sample and analyzed separately.

We further queried the Open Targets Genetics to obtain functional annotations, molecular qualitative trait locus (QTL) evidence, GWAS credible-set information, and enhancer-to-gene predictions for the prioritized variants^48^.

#### Sensitivity analysis

Genetic variants found enriched and underrepresented in our MVP sample were also identified in the HUNT-WGS data. The HUNT Study, is the largest population-based health study in Norway, primarily conducted in the northern region of Trøndelag County^49,50^. The region is predominantly rural and its population is fairly representative of Norway regarding socio-demographic characteristics, as well as mortality and morbidity^51^. Low-coverage (5x) WGS data of 2201 HUNT participants mapped to GRCh37/hg19 reference genome were used as an independent population-based comparison dataset.

Details of the HUNT-WGS sequencing, data processing, and quality control procedures have been described previously^40^. Allele counts were compared between gnomAD non-Finnish European controls and HUNT-WGS cohort and subsequently between our MVP sample (n = 92) and HUNTWGS cohort. In order to harmonize with HUNT-WGS data, *LiftOver* from UCSC Genome Browser was used to convert the genomic positions of the genetic variants from GRCh38/hg38 to GRCh37/hg19^52^. Fisher’s exact test was applied with Bonferroni correction to account for multiple testing.

## Results

### Baseline characteristics

We included 93 patients diagnosed with MVP and/or MAD, of which 92 patients had WGS data passed the genomic quality control checks. Detailed clinical information was available for 89 of these patients (median age 55 years (IQR 39-62), 56% females), of which 37 (42%) patients fulfilled the European Heart Rhythm Association (EHRA) diagnostic criteria for AMVP^32^ (Table 1). The mean left ventricular ejection fraction was 55% (IQR 53-61), 9 patients (10%) had myxomatous mitral valve and 80 (90%) had a fibroelastic deficiency. None of the patients in the cohort were diagnosed with a connective tissue disorder.

**Table 1:** Clinical characteristics of patients with MVP and/or MAD.

|  | n = 89 |
| --- | --- |
| Age at inclusion, median years (IQR) | 55 (39-62) |
| Female, n (%) | 50 (56) |
| EHRA, AMVP diagnosis<sup>a</sup>, n (%) | 37 (42) |
| PVC ≥ 5%, n (%) | 12 (32) |
| Complex ventricular arrhythmia, n (%) | 33 (89) |
| **Echocardiography** |  |
| Verified MVP, n (%) | 81 (91) |
| Mitral regurgitation, n (%) | 58 (65) |
| Bileaflet MVP, n (%) | 42 (47) |
| Left ventricular ejection fraction, mean % (IQR) | 55 (53-61) |
| Inferolateral mitral annular disjunction, n (%) | 67 (75) |
| Classical ≥5mm one or more flail, Barlow, n (%) | 9 (10) |

<sup>a</sup>AMVP diagnosis based on European Heart Rhythm Association (EHRA) criteria<sup>32</sup>

### Genetic results

#### Genes related to non-syndromic MVP and cardiomyopathies

Among the 102 119 genetic variants investigated, 789 variants were significantly associated with MVP and/or MAD at the Bonferroni threshold p < 4.9E-7 (Supplementary file, Table S4), of which 14 genetic variants were in coding or regulatory gene regions. After pruning, 111 independent variants were identified as significantly associated (Supplementary file, Table S5), of which 4 genetic variants were in coding or regulatory gene regions of chromosome 12, 16 and X (Table 2).

**Table 2:** Genetic variants after LD pruning in the coding and regulatory regions of genes related to non-syndromic MVP, MVP and cardiomyopathies, and genes having lesser-known associations with MVP that were associated in our analysis at Bonferroni p-value threshold of 4.9E-7.

| Chromosome | Genomic position (GRCh38) | rsID | REF | ALT | Functional annotation | Gene | Exonic consequence | MVP sample |  | gnomAD NFE |  | Fisher's exact test p-value |
| --- | --- | --- | --- | --- | --- | --- | --- | --- | --- | --- | --- | --- |
|  |  |  |  |  |  |  |  | ALT frequency | Homozygous carriers | ALT frequency | Homozygous carriers |  |
| chr12 | 114355480 | rs201961625 | CCT | C | UTR3 | TBX5 | . | 0.03 | 0 | 7.39E-05 | 0 | 3.46E-11 |
| chr16 | 17470801 | rs117041807 | G | C | UTR5 | XYLT1 | . | 0.05 | 1 | 0.19 | 1193 | 6.50E-08 |
| chr16 | 25692305 | rs533546276 | AGCG | A | UTR5 | HS3ST4 | . | 0.09 | 7 | 0.80 | 19223 | 3.82E-96 |
| chrX | 154362755 | rs202109957 | G | T | exonic | FLNA | nonsynonymous SNV | 0.02 | 1 | 5.64E-05 | 0 | 4.75E-09 |

REF = Reference allele; ALT = Alternate allele; NFE = Non-Finnish European ancestry group

A missense variant in the *FLNA* gene resulting in an amino acid substitution (c.2310C>A:p.Asn770Lys) was significantly enriched in our sample. Additionally, three variants located in UTR of the genes *TBX5* (T-box transcription factor 5), *XYLT1*, and *HS3ST4* were associated. Among these, variant rs201961625 in gene *TBX5* (3’ UTR) was significantly enriched in our sample, while variants rs117041807 (*XYLT1*, 5’ UTR) and rs533546276 (*HS3ST4*, 5’ UTR) were significantly underrepresented compared to the controls.

#### Genes related to syndromes

Among the 31 636 genetic variants investigated, 126 variants were significantly associated with MVP and/or MAD at the Bonferroni threshold p < 1.58E-6 (Supplementary file, Table S6), of which three genetic variants were in coding or regulatory gene regions. After pruning, 30 independent variants were significantly associated (Supplementary file, Table S7), of which three genetic variants were in coding or regulatory gene regions of chromosome 5, 7 and 18 (Table 3).

**Table 3:** Genetic variants after LD pruning in the coding and regulatory regions of genes related to syndromic MVP that were associated in our analysis at Bonferroni p-value threshold of 1.58E-6.

| Chromosome | Genomic position (GRCh38) | rsID | REF | ALT | Functional annotation | Gene | Exonic consequence | MVP sample |  | gnomAD NFE |  | Fisher's exact test p-value |
| --- | --- | --- | --- | --- | --- | --- | --- | --- | --- | --- | --- | --- |
|  |  |  |  |  |  |  |  | ALT frequency | Homozygous carriers | ALT frequency | Homozygous carriers |  |
| chr5 | 172454519 | rs112436148 | G | C | UTR5 | *SH3PXD2B* | . | 0.03 | 1 | 0.35 | 3922 | 1.52E-26 |
| chr7 | 94406291 | rs144540908 | T | G | exonic | *COL1A2* | synonymous SNV | 0.02 | 0 | 3.97E-04 | 0 | 1.52E-06 |
| chr18 | 51082732 | rs559471969 | G | GT | UTR3 | *SMAD4* | . | 0.06 | 0 | 1.26E-03 | 0 | 3.54E-15 |

REF = Reference allele; ALT = Alternate allele; NFE = Non-Finnish European ancestry group

A synonymous variant in the *COL1A2* gene was significantly enriched in our sample compared to the controls. Although it does not alter the amino acid sequence (c.582T>G:p.Ala194=), synonymous variants may affect splicing or mRNA stability. Additionally, two UTR variants were identified: rs112436148 in the 5’ UTR of gene *SH3PXD2B* was significantly underrepresented, and rs559471969 in the 3′ UTR of gene *SMAD4* was significantly enriched in our sample, suggesting potential regulatory effects.

#### Genes related to cardiac arrythmias

Among the 573 581 genetic variants investigated, 2511 variants were significantly associated with MVP and/or MAD at the Bonferroni threshold p < 8.72E-8 (Supplementary file, Table S8), of which 39 genetic variants were in coding or regulatory gene regions. After pruning, 451 independent variants were significantly associated (Supplementary file, Table S9), of which 13 genetic variants were in coding or regulatory gene regions of chromosome 1, 2, 3, 4, 5, 6, 12, 14, 20 and 22 (Table 4).

**Table 4:**
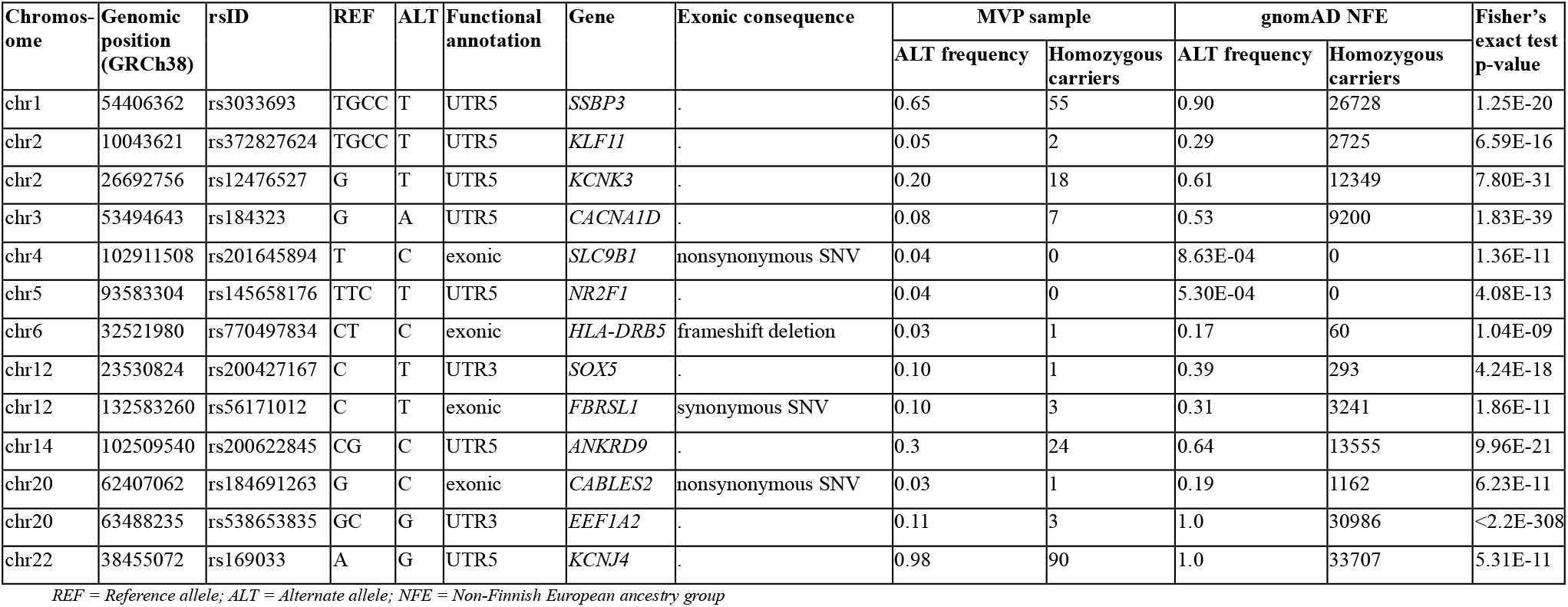
Genetic variants after LD pruning in the coding and regulatory regions of genes related to cardiac arrythmias that were associated in our analysis at Bonferroni p-value threshold of 8.72E-8.

A missense variant in *SLC9B1* (c.859A>G:p.Ile287Val) was significantly enriched in our sample compared to the controls, while three other exonic variants — a missense variant in *CABLES2* (c.215C>G:p.Pro72Arg), a frameshift deletion variant in *HLA-DRB5* (c.294del:p.Asp99fs) and a synonymous variant in *FBRSL1* (c.1870C>T:p.Leu624Leu, c.2620C>T:p.Leu874Leu, c.2491C>T:p.Leu831Leu, c.2545C>T:p.Leu849Leu) — were significantly underrepresented. Furthermore, multiple UTR variants in *SSBP3, KLF11, KCNK3, CACNA1D, NR2F1, SOX5, ANKRD9, EEF1A2* and *KCNJ4* showed strong associations, with only the variant rs145658176 in *NR2F1* significantly enriched and all others underrepresented in our sample, suggesting potential regulatory effects.

#### Sensitivity analysis

In the HUNT-WGS cohort, we had sufficient coverage for variant calling for 82 variants from the list of 789 variants (10.4%) associated with MVP and/or MAD in genes related to non-syndromic MVP and cardiomyopathies (Supplementary file, Table S10). We also identified genotypes for 15 out of 126 variants (11.9%) and 258 out of 2511 variants (10.3%) associated with MVP and/or MAD in genes related to syndromic MVP and cardiac arrythmias, respectively (Supplementary file, Table S11 and S12). For a subset of variants that differed significantly between our sample and gnomAD non-Finnish European controls, significant differences were also observed between our sample and the HUNTWGS cohort, whereas no significant differences were detected between HUNT-WGS and gnomAD controls. However, other subset of variants demonstrated discordant results across pairwise comparisons, including variants that differed significantly from gnomAD non-Finnish European controls but not from the HUNT-WGS cohort.

## Discussion

This is the first WGS–based targeted genetic analysis in patients with MVP and/or MAD. We systematically investigated genetic variants across curated gene sets informed by prior literature: (i) genes previously linked to non-syndromic MVP, MVP and cardiomyopathies, and those with emerging linkages with MVP; (ii) genes implicated in syndromic forms of MVP and connective tissue disorders; and (iii) genes involved in cardiac arrhythmias. By focusing on these biologically and clinically relevant genomic regions, rather than adopting a hypothesis-free genome-wide approach, we enhance the power to detect true associations while strengthening the interpretability of these findings. A brief summary with findings from the Open Targets Genetics for the prioritized variants is presented in Supplementary file, Table S13.

### Non-syndromic MVP and cardiomyopathic genes

A key finding of our panel-based analysis is the significant enrichment of a missense variant in *FLNA* (rs202109957) in MVP cases, having direct protein-altering effects (N770; c.2310C>A:p.Asn770Lys). This aligns with the known role of *FLNA*. Previous studies have implicated mutations in this gene as causative in myxomatous mitral valve dystrophy^26,27^, highlighting its function in valvular development and maintenance of the cardiac extracellular matrix (ECM)^53^. Our finding, observed within a nonsyndromic MVP cohort, provides further support for *FLNA* as a central component of non-syndromic MVP risk, consistent with targeted sequencing data from family studies^26,27^.

Additionally, we found significant associations for regulatory variants within the UTRs of genes *TBX5, XYLT1*, and *HS3ST4*. The 3’ UTR variant in *TBX5* (rs201961625) was enriched in MVP cases. *TBX5* functions as a critical transcription factor that regulates gene expression to direct the normal embryonic development and structural formation of both the heart and upper limbs^54^. While pathogenic *TBX5* variants are classically linked to Holt-Oram syndrome^54^, recent data suggest that subtle regulatory alterations may influence cardiac development and valvular morphogenesis as well^55,56^. Our finding extends this evidence, proposing a possible regulatory effect in MVP not explained by overt loss-of-function mutations^57^. By contrast, underrepresented variants in *XYLT1* (rs117041807) and *HS3ST4* (rs533546276) may reflect protective roles or allelic heterogeneity at these loci. The Open Targets Genetics highlighted that 5’ UTR variant rs117041807 in *XYLT1* is supported by enhancer-to-gene predictions linking the locus to *XYLT1* across multiple tissues, including kidney, dorsolateral prefrontal cortex, spleen, testis and brain microvascular endothelial cells, as well as expression QTL (eQTL) in immune cells and skin. Both *XYLT1* and *HS3ST4* genes participate in glycosaminoglycan or heparan sulfate biosynthesis—biological processes relevant to valve structure and susceptibility to degenerative changes^58,59^. These findings warrant functional follow-up to elucidate how these noncoding variants might modulate MVP risk, especially in the context of the ECM remodeling characteristic of MVP.

### Syndromic genes

Within the gene set implicated in syndromic MVP and collagenopathies, we observed a synonymous variant in *COL1A2* (c.582T>G:p.Ala194=) to be significantly enriched in MVP. Despite no change in amino acid sequence, such variants may exert functional effects by modulating the regulation of gene expression at multiple levels, including mRNA stability, splicing, or translation^57^. *COL1A2* encodes the *α2* chain of type I collagen, a major structural component of the ECM in cardiac valves and other connective tissues. Pathogenic variants in this gene are well known in osteogenesis imperfecta and Ehlers-Danlos syndrome^14,15^. Our finding aligns with the hypothesis that subtle genetic variation in collagen-encoding genes contributes to MVP through ECM dysregulation or altered valve biomechanics.

Two additional variants mapped to UTRs were found associated to MVP cases, suggestive of regulatory effects. The 5’ UTR variant rs112436148 in *SH3PXD2B* was significantly underrepresented in MVP cases, potentially indicating a protective role or reflecting allelic heterogeneity at this locus. This variant is supported by eQTL signals in monocytes and associated with uncertain significance to Frank-Ter Haar syndrome or Dermato-cardio-skeletal syndrome–Borrone type as highlighted in the Open Targets Genetics. The *SH3PXD2B* gene encodes a protein involved in cytoskeletal reorganization and ECM degradation through regulation of podosome formation^60^. Disruptions in matrix remodeling pathways may influence valve structure and susceptibility to prolapse, although direct involvement of *SH3PXD2B* in MVP has not been extensively documented, warranting further functional validation. Notably, the 3’ UTR variant rs559471969 in *SMAD4* was significantly enriched in MVP cases. *SMAD4* is a pivotal mediator of TGF-β signaling pathway, which orchestrates cellular processes vital for cardiac development and ECM homeostasis^61^. Dysregulated TGF-β signaling has been implicated in several connective tissue disorders with valvular involvement, such as Marfan syndrome and Loeys-Dietz syndrome^11,12^. Supporting these findings, a previous clinical study showed high prevalence of MVP and MAD in patients with Marfan syndrome and Loeys-Dietz syndrome^13^. Variants affecting regulatory regions of *SMAD4* could influence gene expression profiles during valvulogenesis or in maintaining valve integrity, thus contributing to MVP pathophysiology outside classical syndromic presentations.

### Arrhythmia-related genes and arrhythmic substrate in MVP

Among the exonic variants, the missense variant in *SLC9B1* (c.859A>G:p.Ile287Val) was significantly enriched in cases. Although *SLC9B1* is primarily known as a sodium/hydrogen exchanger involved in cellular ion homeostasis^62^, its role in cardiac electrophysiology is not extensively characterized. The enrichment of this variant could indicate possible novel contributions to arrhythmic vulnerability or myocardial cell function influencing MVP phenotype, warranting further investigation.

Conversely, three exonic variants were significantly underrepresented in MVP cases – a missense variant in *CABLES2* (c.215C>G:p.Pro72Arg), a frameshift deletion variant in *HLA-DRB5* (c.294del:p.Asp99fs), and a synonymous variant in *FBRSL1* (c.1870C>T:p.Leu624Leu, c.2620C>T:p.Leu874Leu, c.2491C>T:p.Leu831Leu, c.2545C>T:p.Leu849Leu). This missense variant rs184691263 in *CABLES2* is in an enhancer region that regulates *CABLES2* across multiple tissues, including heart, lung, spleen, placenta, skin, and CD4+ T cells, as well as overlaps with GWAS credible sets for QT interval and lean body mass/body composition traits as highlighted in the Open Targets Genetics. *CABLES2* has been implicated in cell cycle regulation and apoptosis^63^, and while its direct role in cardiac electrophysiology remains unclear, this finding raises the possibility of protective or modifying effects on arrhythmogenic pathways related to MVP. The frameshift variant in *HLA-DRB5*, part of the major histocompatibility complex locus with roles in immune regulation, may indirectly influence myocardial inflammation or remodeling, both contributors to arrhythmogenesis^64^. Although developmental studies of truncating *FBRSL1* (fibrosin-like 1) variants in animal models show clear effects on heart formation^65^, the relevance of synonymous variant in *FBRSL1* is less evident; however, it might still be having regulatory or splicing effects altering protein expression^57^, highlighting the need for further functional investigation.

Numerous variants in the UTRs of key arrhythmia-relevant genes with established or putative roles in cardiac electrophysiology and structure, including *SSBP3, KLF11, KCNK3, CACNA1D, NR2F1, SOX5, ANKRD9, EEF1A*2, and *KCNJ4* were found associated with MVP. The Open Targets Genetics highlighted that 5’ UTR variant rs12476527 in *KCNK3* overlaps with GWAS credible sets for hypertension/blood pressure phenotypes and rs184323 in *CACNA1D* overlaps with GWAS credible sets for height/body composition related traits. Similarly, the 5’ UTR variant rs200622845 in *ANKRD9* showed strong support, with convergent eQTL, spliceQTL, and transcript-usage QTL evidence in skeletal muscle, adipose tissue and skin, as well as overlaps with GWAS credible sets for QT interval, JT interval, kidney function, and erythrocyte traits. Notably, all but one—rs145658176 in *NR2F1*— were underrepresented in MVP cases, suggesting a complex landscape where certain regulatory alleles may confer protective effects or reflect population-specific haplotype structures. The enrichment of the UTR variant in *NR2F1*, a nuclear receptor involved in transcriptional regulation, suggests a possible influence on cardiac gene networks affecting arrhythmic vulnerability^66^. Although its role remains incompletely defined, *NR2F1* is expressed in cardiac tissue, indicating a potential involvement in cardiac development and structural integrity^66^. Given its role in transcriptional regulation, variants in *NR2F1* may alter gene expression during valve morphogenesis, thereby leading mitral valve abnormalities.

*KCNK3* and *KCNJ4* encode potassium channels that contribute to myocardial repolarization; alterations in their expression or function may predispose to arrhythmias through modulation of action potential duration and refractoriness^67,68^. Similarly, variants in *CACNA1D*, encoding the L-type calcium channel *α1D* subunit, implicate calcium handling abnormalities as arrhythmogenic contributors^69^. Other genes, such as *SSBP3* and *KLF11*, have roles in DNA binding and transcriptional control with emerging but not fully mapped cardiac functions^70,71^. The significant underrepresentation of variants in these loci could indicate altered gene regulation or evolutionary constraints in MVP patients.

### Limitations

Our approach—targeting predefined gene sets informed by prior research—enhances the power and interpretive value of the observed associations, but omits potentially relevant loci outside these regions. However, restricting the number of potential targets is necessary when the sample is limited (n = 92), in order for true positives to survive multiple testing correction.

As with all candidate gene studies, our findings are bounded by the current understanding of gene-phenotype relationships. Functional annotation, particularly of non-coding variants, remains limited, and the observed associations require experimental validation. Genetic heterogeneity, population stratification, and the absence of whole-genome interrogation are further considerations.

Overall, MVP and/or MAD associated variants identified in our small patient sample limited the ability to detect statistically significant differences in allele frequencies. Moreover, the use of gnomAD as source for the control population could possibly lead to differences between cases and controls that are due to allele frequency differences between genetic ancestry groups (i.e., Norwegians versus non-Finnish Europeans), or because of different protocols in determining variant frequencies. To address this potential limitation, we performed a sensitivity analysis using the Norwegian HUNT-WGS cohort as an ancestry-matched reference population. For many variants, allele frequency differences observed relative to gnomAD non-Finnish European controls remained significant when compared with HUNTWGS cohort, while no significant differences were observed between HUNT-WGS and gnomAD non-Finnish Europeans, suggesting that our findings are unlikely to be explained solely by population stratification. Nevertheless, a good proportion of variants yielded discrepant results, including variants that differed from gnomAD non-Finnish Europeans but not from HUNT-WGS cohort, indicating that some findings may reflect population-specific allele frequency variation rather than disease association. In addition, a few variants differed significantly across all comparisons, suggesting that although population structure may contribute to allele frequency differences, it cannot fully account for the observed enrichment or underrepresentation in our sample. This underscores the importance of interpreting variant enrichment or underrepresentation cautiously, particularly when variant data are absent or sparse in reference cohort derived from the matched ancestry, i.e. HUNT-WGS cohort data having low-coverage (5x). These discrepant findings may also reflect characteristics of the HUNTWGS cohort, which represents a population-based sample randomly selected from HUNT participants rather than a cohort of healthy controls. Although only a small proportion of HUNT participants would be expected to have the outcome of interest, differences in cohort composition, and variant calling protocols may nevertheless contribute to differences in observed allele frequencies.

### Implications and future directions

Our study confirms and extends existing evidence implicating specific genes in MVP, underscoring the contribution of both coding and regulatory variation to heritable risk. The observed associations provide valuable leads for future functional genomics and may ultimately support personalized risk prediction for patients with MVP. Replication in larger, independent cohorts with diverse ancestries, together with integrative analyses of transcriptomic, proteomic, and clinical data, will be essential to ensure robust generalization and to elucidate the mechanisms through which the implicated variants contribute to valve pathology.

The identification of associated variants in genes connected with connective tissue and syndromic MVP highlights the importance of ECM and TGF-β signaling pathways in MVP susceptibility. While a part of our analysis focused on variants in a targeted set of syndromic MVP genes, the results indicate that inherited variation within these loci may also contribute to non-syndromic MVP phenotypes or modulate disease severity and progression. This supports the view that MVP represents a genetically heterogeneous condition with overlapping molecular mechanisms between non-syndromic and syndromic forms. Another part of our analysis identified variants in arrhythmia-related genes—both coding and regulatory—might contribute to an inherited substrate that interacts with biomechanical stresses on the mitral valve to increase arrhythmia susceptibility. Collectively, our findings add to the growing recognition that MVP extends beyond a purely valvular abnormality, encompassing myocardial and electrophysiological remodeling processes that heighten the risk of life-threatening arrhythmias and SCD.

Future research will benefit from integrating these variants into polygenic risk models, leveraging functional genomics to dissect regulatory and coding effects, and pursuing prospective clinical studies to evaluate implications for risk stratification, management, and possible therapeutic targeting.

## Conclusion

This candidate gene study expands the genetic landscape of MVP and/or MAD by demonstrating robust associations involving *FLNA, TBX5, XYLT1, HS3ST4, COL1A2, SH3PXD2B* and *SMAD4*, as well as numerous genes implicated in arrhythmogenesis – *SLC9B1, CABLES2, HLA-DRB5, FBRSL1, SSBP3, KLF11, KCNK3, CACNA1D, NR2F1, SOX5, ANKRD9, EEF1A2* and *KCNJ4*. By integrating coding and regulatory genetic data across non-syndromic and syndromic MVP loci, and arrhythmia-associated genes, we highlight the complex, multifactorial genetic architecture underlying MVP. Our findings substantially advance our understanding of the genetic determinants of MVP and/or MAD, lay the groundwork for future studies and may inform the development of genetically guided stratification strategies in patients with MVP and/or MAD.

## Data Availability

All data produced in the present study can be available upon reasonable request to the authors

https://gnomad.broadinstitute.org/data

## Acknowledgements

NA, JB, CRN, CF, CB, AIC, NEH, EC, KH, and KHH, are affiliated to the Precision Health Center for Optimized Cardiac Care (ProCardio) funded by the Norwegian Research Council (#309762).

BNW is supported by the European Union’s Horizon Europe Research and Innovation Programme under the Marie Skłodowska-Curie grant agreement number 101110878.

